# Thermal Camera Detection of High Temperature for Mass COVID Screening

**DOI:** 10.1101/2021.05.05.21256285

**Authors:** Richard S. Maguire, Matt Hogg, Iain D. Carrie, Maria Blaney, Antonin Couturier, Lucy Longbottom, Jack Thomson, Nicola Baxter, Aimee Thompson, Craig Warren, David J. Lowe

## Abstract

The COVID-19 [SARS-COV-2] pandemic has had a devastating global impact, with both the human and socio economic costs being severe. One result of the COVID-19 pandemic is the emergence of an urgent requirement for effective techniques and technologies for screening individuals showing symptoms of infection in a non-invasive and non-contact way. Systems that exploit thermal imaging technology to screen individuals show promise to satisfy the desired criteria, including offering a non-contact, non-invasive method of temperature measurement. Furthermore, the potential for mass and passive screening makes thermal imaging systems an attractive technology where current ‘standard of care’ methods are not practical.

Critically, any fever screening solution must be capable of accurate temperature measurement and subsequent prediction of core temperature. This is essential to ensure a high sensitivity in identifying fever while maintaining a low rate of false positives. This paper discusses the results and analysis of a clinical trial undertaken by Thales UK Ltd and the Queen Elizabeth University Teaching Hospital in Glasgow to assess the accuracy and operation of the High Temperature Detection (HTD) system developed by Thales UK Ltd when used in a clinical setting.

Results of this single centre prospective observational cohort study show that the measured laboratory accuracy of the Thales HTD system (***RMSE***=**0**.**1**°C) is comparable to the accuracy when used in a clinical setting (***RMSE*** = **0**.**1**5°C) when measuring a calibrated blackbody source at typical skin temperature. For measurement of forehead skin temperature, the system produced results commensurate with close contact measurement methods (***R*** = **0**.**86, *Mean error***=**0**.**05**°***C***).. Compared to measured tympanic temperatures, measurement of the forehead skin temperature by the HTD system showed a moderate correlation (***R*** = **0**.**43**),), which is stronger than close contact IR forehead thermometers **(*R*** = **0**.**20**,**0**.**35**) An improved correlation was observed between the maximum facial temperature measured by the HTD system and measured tympanic temperatures (***R*** = **0**.**53**), which is significantly stronger than the close contact methods. A linear predictive model for tympanic temperature based on the measured maximum facial temperatures resulted in a root mean square error (***RMSE*** = **0**.**50**°C) that is marginally larger than what is expected as a compound of errors in the measuring devices (***RMSE***=**0**.**45**°***C***).

The study demonstrates that the HTD could be applied in the clinical and non-clinical setting as a screening mechanism to detect citizens with raised temperature. This approach would enable high volume surveillance and identification of individuals that contribute to further spread of COVID-19. Deployment of the HTD system could be implemented as part of a screening tool to support measures to enhance public safety and confidence in areas of high throughput, such as airports, shopping centres or places of work.

## I. Introduction

The COVID-19 [SARS-COV-2] pandemic has had devastating consequences globally since identification in late December 2019, with both the human and socio-economic costs being severe [1]. While the majority of COVID-19 cases are asymptomatic or have limited symptoms there is a proportion that develop severe respiratory illness leading to critical illness [2]. Due to the high mortality rate, lack of effective treatment and highly transmissible nature of the virus, there is a need to limit the spread of the virus by developing robust measures to rapidly screen and test individuals [3]. Diagnosis of COVID-19 continues to rely on ‘gold standard’ high performance laboratory based PCR testing but there is increasingly availability of point of care tests with improving diagnostic capability [4]. The mainstay of any testing approach relies on identifying the ‘at risk’ individuals either by reporting of symptoms or contact tracing [5]. The case definition of symptoms of COVID-19 have been well described with the WHO recommending testing of those that exhibit (1) a new continuous cough, (2) a fever (3) a loss/change in smell or taste [6]. One area of significant research is the identification of individuals exhibiting a fever as a symptom with the reported incidence varying [7]. Guan et al, in an analysis of 1099 patients with laboratory confirmed COVID-19, observed that a fever was recorded in 43.8% of cases on admission to hospital and in 88.7% of cases during hospitalisation [8]. Another study by Ishikawa et al reported similar results with 50% exhibiting fever on admission and 78.5% during hospitalisation in a study of 7614 patients [9].

Due to the high prevalence of fever in COVID-19 cases there is a strong worldwide desire to develop effective mass screening techniques which can quickly and accurately determine if an individual is exhibiting a fever [10] [11]. An ideal solution would be minimally invasive, with little discomfort to the individual, would minimise contact with the operator while providing accurate and reliable temperature measurements. Furthermore, a solution should have minimal size and cost and ideally be capable of mass screening. Thermal cameras provide a non-invasive, non-contact solution for fever screening and offer the opportunity to deliver a mass screening solution to augment public health strategies.

Thermal cameras have the potential to be deployed to measure subject’s temperature to predict core temperature to enable rapid identification of febrile subjects. It is important to note that skin temperature and core body temperature are not equivalent and that the skin temperature changes in response to a number of environmental conditions to try and maintain a consistent core body temperature [12]. Whilst the meta-analysis of earlier research by Tipton et al [12] showed that there was poor correlation between measured skin temperature and core body temperature, more recent research in this area driven by improvements in uncooled thermal imaging technology over the last decade show significantly improved correlation between maximum facial skin temperature and core body temperature measured orally [13]. In addition to this, operation of the system in a controlled environment, for example a transport security setting, could reduce the variability in some of the environmental conditions described by Tipton et al [12].

One of the main drawbacks for commercially available infrared camera systems for measuring skin temperature is that the majority of systems have a typical off the shelf laboratory accuracy of ±0.5°C. For systems that incorporate an in scene blackbody source for calibration, this can typically be improved to ±0.3°C, however this requirement increases cost, size and reduces the flexibility of the system. In order to minimise size, cost and complexity, an ideal candidate system would not require an external blackbody source. In addition to this, accuracies greater than ±0.3°C would be critical in ensuring high sensitivity of the fever screening system while minimising false positives.

## II. Study Aims

This study had three key aims from the outset which are described below:

- Assess the temperature measurement accuracy of a thermal imager based screening system in a clinical setting, and to compare against current ‘standard of care’ methods of determining core temperature.
- Ascertain the correlation between skin temperature measurement and core body temperature measured using a tympanic thermometer.
- Developing and proposing a predictive model to predict core temperature based on measured skin temperature.

## III. Study Methods

This single centre prospective observational cohort study was performed at the Queen Elizabeth University Hospital Emergency Department, Glasgow. Ethical approval was granted by Camden & Kings Cross Research and Ethics Committee (20/HRA/4413). The study was registered on clinicaltrials.gov NCT04792450. The study ran for four weeks from 17/11/2020 to 16/12/2020.

Two cohorts were recruited for testing, a patient and a staff group. Inclusion criteria were patients over the age of 16 years old, able to understand and read English and able to give informed consent. Recruitment took place when research staff were available and was from 0800 to 2000 and was a convenience sample. Patients or staff were invited to review the patient information sheet and following this consented to participate, with all measurements being conducted in the well-controlled ambient environment of the hospital (temperature, humidity and indoor lighting) controlling many of the environmental concerns raised by Tipton et al [12]. Researchers were trained by Thales technical staff on the use of the camera by developing a set of standard operating procedures and through video call. Personal Protection Equipment in accordance with NHS Greater Glasgow and Clyde was adhered to.

### A. Hardware Description

Thales in Glasgow working in collaboration with the Queen Elizabeth University Hospital have developed a thermal imaging system (called the High Temperature Detection (HTD) Camera system shown in Figure 1). The system is capable of performing temperature measurement of subjects at range, without the need for an external blackbody calibration source. In addition to this, proprietary facial detection and tracking enables fast, easy measurement and contact-free operation.

**Figure 1.**
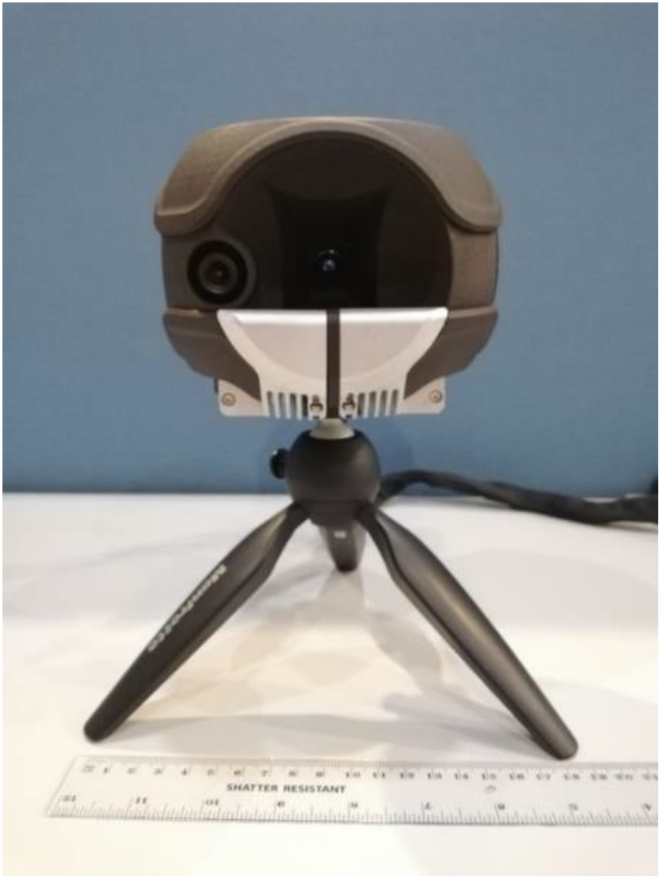
Thales HTD Camera System

To assess the accuracy of the HTD system and measure agreement with other current ‘standard of care’ methods, the following additional hardware was required:

- Engineering grade infra-red thermometer – this is a calibrated infrared thermometer designed for measuring the temperature of a wide range of surfaces with a measurement accuracy after calibration of ±1% for skin temperature; this generally equates to approximately ±0.35°C.
- Commercial off the shelf forehead thermometer – this is a commercially available forehead thermometer which measures the skin temperature of the subject and then applies a proprietary adjustment to estimate core temperature, with a claimed accuracy of ±0.2°C.
- Clinical tympanic thermometer – this is a clinical thermometer used to measure temperature of patients in the emergency department with an accuracy of ±0.3°C after calibration.
- A test tablet PC with associated software was supplied to capture imagery and data from the HTD camera system and all the thermometers listed above.
- A calibrated blackbody source with a set point accuracy of ±0.03°C.

### B. Data Gathering Description

The HTD camera and reference thermometers were used by clinicians in the hospital Emergency Department (ED) to measure temperatures of 100 staff, 100 patients and a calibrated blackbody source. Sample size was chosen to give a confidence interval of ±0.34σ on the limits of agreement for each sub-cohort and a confidence interval of ±0.24σ on the full cohort [14]. A demographic breakdown of the study participants is given in Table 1, it is worth noting that two entries were recorded for all participants. All collected data comparing the HTD system to other temperature measurements was analysed anonymously without knowledge of age; sex or ethnicity to minimise the potential for bias in the analysis and conclusions.

**Table 1.**
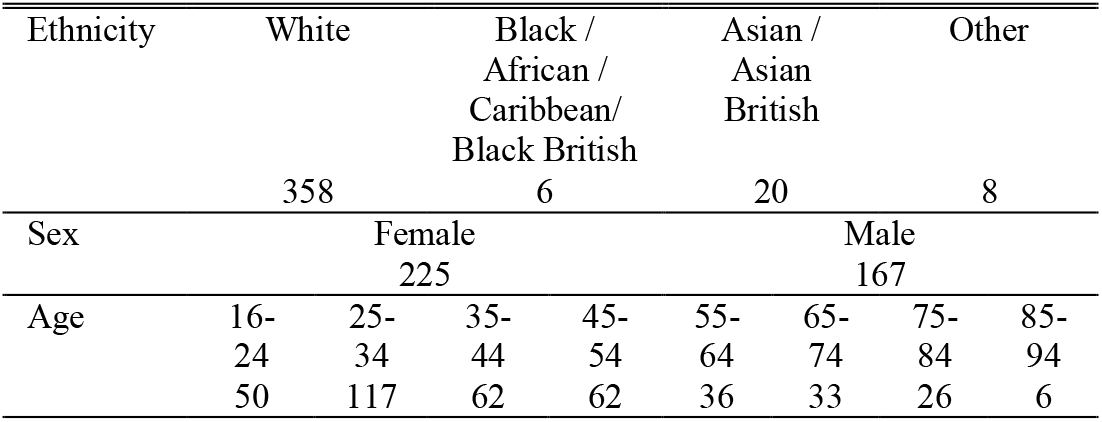
Sample size by demographic

Measurements of staff were recorded in a staff training room while measurements of patients were recorded in the ED. Throughout the experiment, dedicated measurement of the blackbody by both the HTD camera and the reference thermometers was performed to identify any drift of accuracy in any of the measuring instruments. Furthermore, the blackbody was positioned so that it was visible in the captured images for staff recordings only. This was to provide further assurance in the accuracy of the system outside of the dedicated blackbody measurements and was not required for the operation of the HTD system. To ensure reliability of tympanic measurements, the Tympanic thermometer was calibrated weekly by the Medical Physics department during the course of the study following manufacturer’s instructions.

In this experiment, all HTD camera measurements were performed at a fixed range of 1.75m. For staff and blackbody measurements this was achieved by setting up a single station for the duration of the experiment. For patients, recordings were taken at a range as close to the predefined range as was possible.

A bespoke test application was used to record temperature measurements and images, in addition to age, sex and ethnicity of the subjects. Recorded temperature measurements were used to measure the accuracy of the HTD camera and assess agreement with the reference thermometers. The recorded images enabled post processing and exploratory studies and were not used in the accuracy assessment of the system.

#### 1) Accuracy of HTD Temperature Measurement in a Laboratory Setting

In advance of the study, the HTD system was measured extensively in a lab environment against a calibrated blackbody source. In all of these tests the HTD camera was positioned in close proximity to the blackbody source such that it was filling the field of view of the camera, this results in a distance of between 5cm and 10cm, between the surface of the blackbody source and the HTD camera system. Three calibration tests were conducted at 36°C, 34°C & 38°C with the temperature of the blackbody source measured by the HTD camera system once every second for an extended period of time (at least 30 minutes for 34° & 38°C and in excess of 1 hour for 36°C). Results of these tests are presented in Section IV.A.

#### 2) Temperature Measurement of a Calibrated Blackbody Source in a Clinical Setting

During the study, the temperature of a calibrated blackbody source was measured by the HTD camera and recorded along with temperatures measured by the 3 reference thermometers. Measurements using the HTD camera were recorded by selecting a region in the image corresponding to the blackbody in the image displayed in the application. For the Engineering and Forehead thermometers, a measurement of the blackbody at the proximity recommended for each product was recorded. For the Tympanic thermometer, a measurement at a distance in the order of 1cm to the blackbody was recorded. It was accepted that for the latter, any specific requirements for proximity required by the product are harder to satisfy when using the instrument in this context compared to its usage in-ear. The blackbody was set to 35°C for all measurements.

#### 3) Temperature Measurement of Patients and Staff in a Clinical Setting

Similarly to the blackbody source, measurements from the HTD camera as well as all 3 reference thermometers were recorded for each member of staff and each patient. For the majority of staff, and a subset of patients, 2 recordings were taken; measurements with mask and/or glasses and measurements without. Furthermore, for subjects where a face was detected by the HTD camera, the thermal statistics of the region spanning the face were also recorded.

## IV. Data Analysis and Results

### A. Accuracy of HTD Temperature Measurement in a Laboratory Setting

Prior to analysing the accuracy of the HTD camera in the clinical setting, results indicating accuracy of the camera in the laboratory setting are presented.

The summary statistics for all measurements are shown in Table 2 below, whilst Figure 2, Figure 3 and Figure 4 shows the error as a yield plot (the percentage of measurements with an absolute error equal to or less than a given value). These results show that, in a laboratory setting, a typical RMSE of <0.1°C is achieved without the requirement of a reference blackbody for infield calibration. Calibration of the blackbody used in these tests indicated that this has an intrinsic error ±0.03°C which is another source of error in the results presented here.

**Table 2:** Summary Statistics for Laboratory Accuracy Tests of HTD System

| Measurement | Mean [$^{\circ}\text{C}$] | Standard Deviation [$^{\circ}\text{C}$] | RMS Error [$^{\circ}\text{C}$] |
| --- | --- | --- | --- |
| 36°C | 36.016 | 0.095 | 0.095 |
| 34°C | 34.003 | 0.090 | 0.090 |
| 38°C | 38.018 | 0.093 | 0.094 |

**Figure 2:**
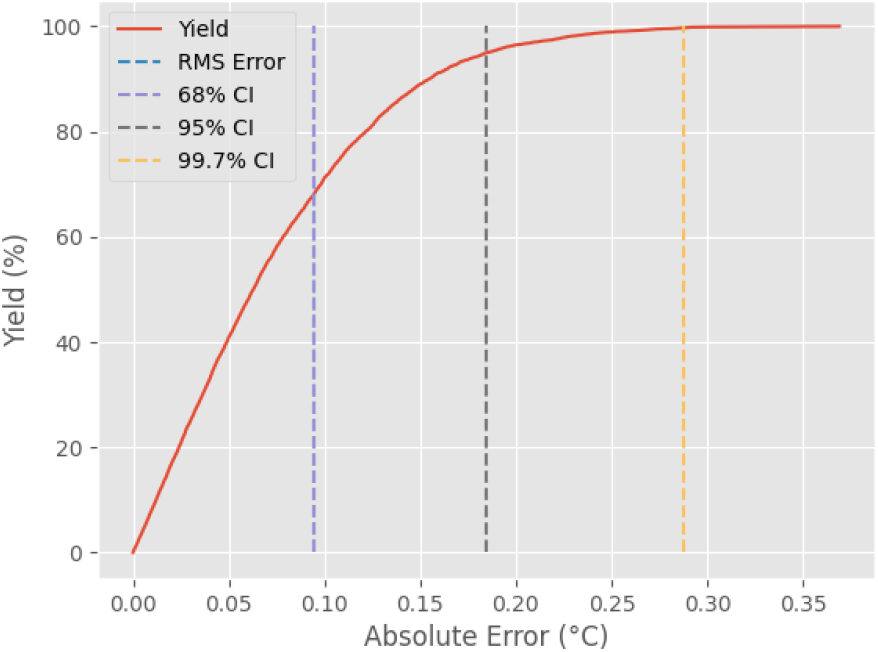
HTD Blackbody Laboratory Accuracy Test at 36°C Yield Analysis

**Figure 3:**
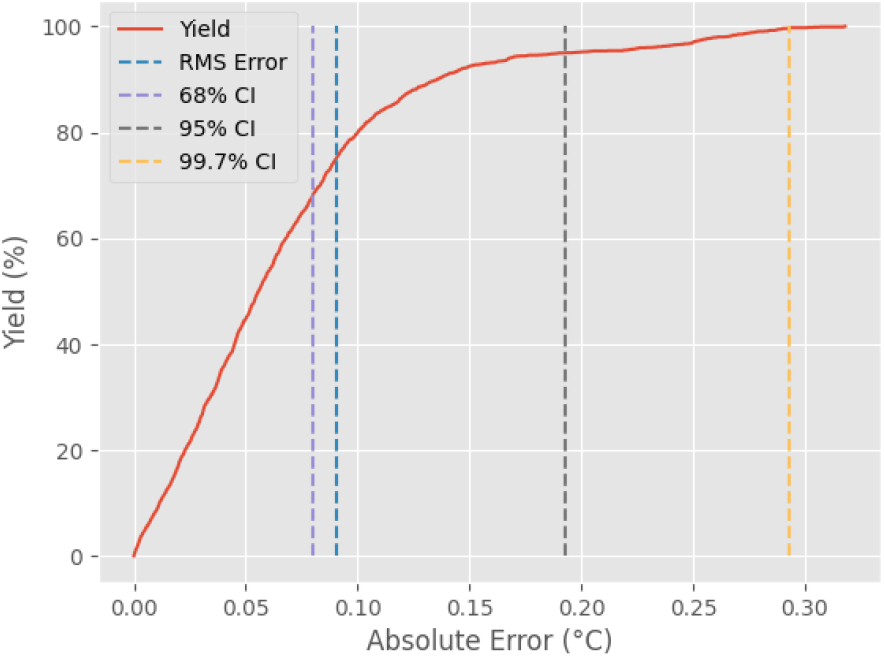
HTD Blackbody Laboratory Accuracy Test at 34°C Yield Analysis

**Figure 4:**
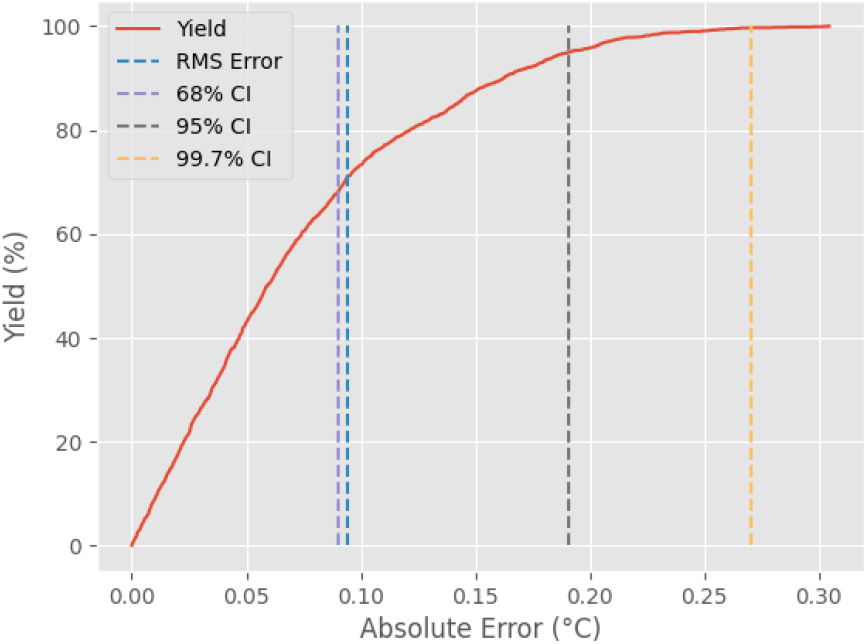
HTD Blackbody Laboratory Accuracy Test at 38°C Yield Analysis

### B. Accuracy of HTD Temperature Measurement in a Clinical Setting

Following completion of the trial, the data was manually cleaned to remove obviously erroneous points (such as those where a blackbody entry was recorded for a patient or thermometer readings were transposed). Furthermore, metadata that was not recorded at capture, for example the identification of the entry as staff or patient was also appended to the data at this stage to allow for analysis of subsets. For a complete list of removed/corrected data points and additional metadata see

Appendix A Data Filtering and Pre-processing.

#### 1) Temperature Measurement of a Calibrated Blackbody Source in a Clinical Setting

Figure 5 shows the distribution of temperature measurements of a calibrated blackbody source for all four instruments when measured by clinical staff, in a clinical setting. For this test, the blackbody source was set to 35°C. Table 3 below shows the summary statistics for each of the four measuring instruments when compared to the blackbody source. In Figure 5 there are five distinct markings; the distribution of the data is shown as a transparent histogram, and in the middle of the distribution there are two horizontal markings showing both the median and mean of the measurements respectively (note that they might be overlaid in some distributions). Finally there are horizontal markings at either end of the distribution and these show the extrema of the measurements.

**Figure 5:**
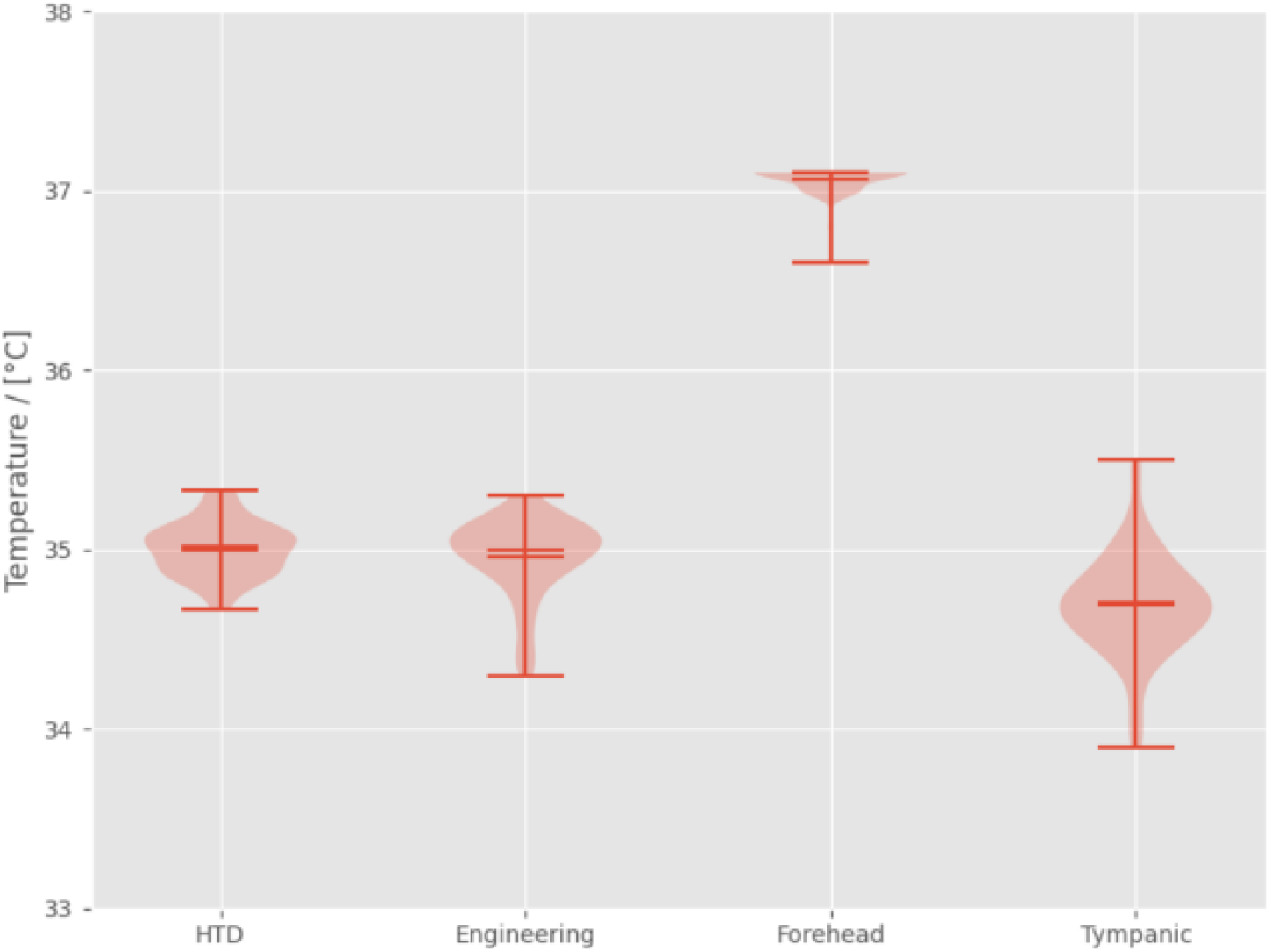
Distribution of Measurements of Calibrated Blackbody Source at 35.0°C

**Table 3:** Summary Statistics for Each Instrument when Measuring a Calibrated Blackbody Source at 35°C

| Instrument | Mean [°C] | Standard Deviation [°C] | RMS Error [°C] |
| --- | --- | --- | --- |
| HTD (at 1.75m) | 35.001 | 0.149 | 0.149 |
| Engineering | 34.955 | 0.217 | 0.221 |
| COTS Forehead | 37.059 | 0.085 | 2.06 |
| Tympanic | 34.689 | 0.283 | 0.423 |

It is presumed that the observed error for each measurement for all instruments is attributable to three error sources: error in the blackbody temperature (provided in Section III.A), intrinsic error of the instrument and operational error. The RMS value for the Engineering IR thermometer is within the ±0.3°C error bounds specified for the thermometer.

The RMS error for the HTD system is within ±0.15°C, which is consistent with the laboratory measurements presented in Section A, with a slight increase in error expected when operated outside of a controlled environment.

The large RMS error of the COTS Forehead thermometer and the offset in the mean value from the blackbody set point is attributed to the core temperature estimation offset applied by the COTS Forehead thermometer. A core temperature estimation algorithm could also result in a reduced standard deviation if this correction were to map a wider distribution of skin temperatures to a smaller distribution of predicted core temperatures; which could explain the small standard deviation measured here.

The RMS value for the tympanic thermometer is larger than the ±0.3°C error bounds specified. This could be attributed to the larger operational error due to the method of measurement of the blackbody being inconsistent to how the thermometer will be used in normal operation (it is worth noting however that the Tympanic thermometer operating manual states that the device should be calibrated against a black body source, this blackbody source is incorporated into a calibration device which sets the thermometer into a calibration mode and automatically calibrates the device). This is in contrast to the COTS Forehead and Engineering thermometer for which the usage is comparable.

Comparison of the RMS error values for the HTD measurement and that of the reference thermometers would indicate that a measurement of skin temperature using the HTD camera would yield a higher accuracy than if it were measured using the Engineering IR thermometer. Due to the core temperature estimation applied by the COTS Forehead thermometer, an exact comparison using the results at a single temperature set point is difficult as no reverse operation was performed on the recorded data. Comparison across a range of actual skin temperatures is left for Section 2)ii). Likewise, a direct comparison between the HTD camera and tympanic thermometer is excluded due to the difference in usage. Results for the tympanic thermometer included here are later referred to when relating HTD measurements to core temperature in Section C.

#### 2) Measurement of Skin Temperature in a Clinical Setting

To understand the performance of the HTD camera in measuring actual skin temperature the forehead measurement from the HTD camera is compared to the Engineering and COTS Forehead thermometers for human entries. In addition to the analysis of the full set of entries, the results are further divided into staff and patient subsets.

##### i) Comparison HTD and Engineering Thermometer

Figure 6 presents the results for comparison of the HTD camera and Engineering thermometer forehead measurements. Results for the three subsets are summarised in Table 4. In Figure 6, the plots on the left show the temperature recorded by the Engineering thermometer plotted against the mean forehead temperature recorded by the HTD system for all study participants and then staff and patients respectively. The plots on the right show the Bland-Altman analysis [15] between the two measurement methods. This analysis is the preferred approach for assessing agreement of two methods of measurement as it evaluates any bias between the measurements as well as estimates an agreement interval. The former is clearly presented in a Bland-Altman plot as the mean difference, while the latter is interpreted from the 95% confidence interval (±1.96 standard deviations). This confidence interval is indicated on the plot as a lower and upper bound, within which, 95% of the differences between the two measurements fall.

**Figure 6:**
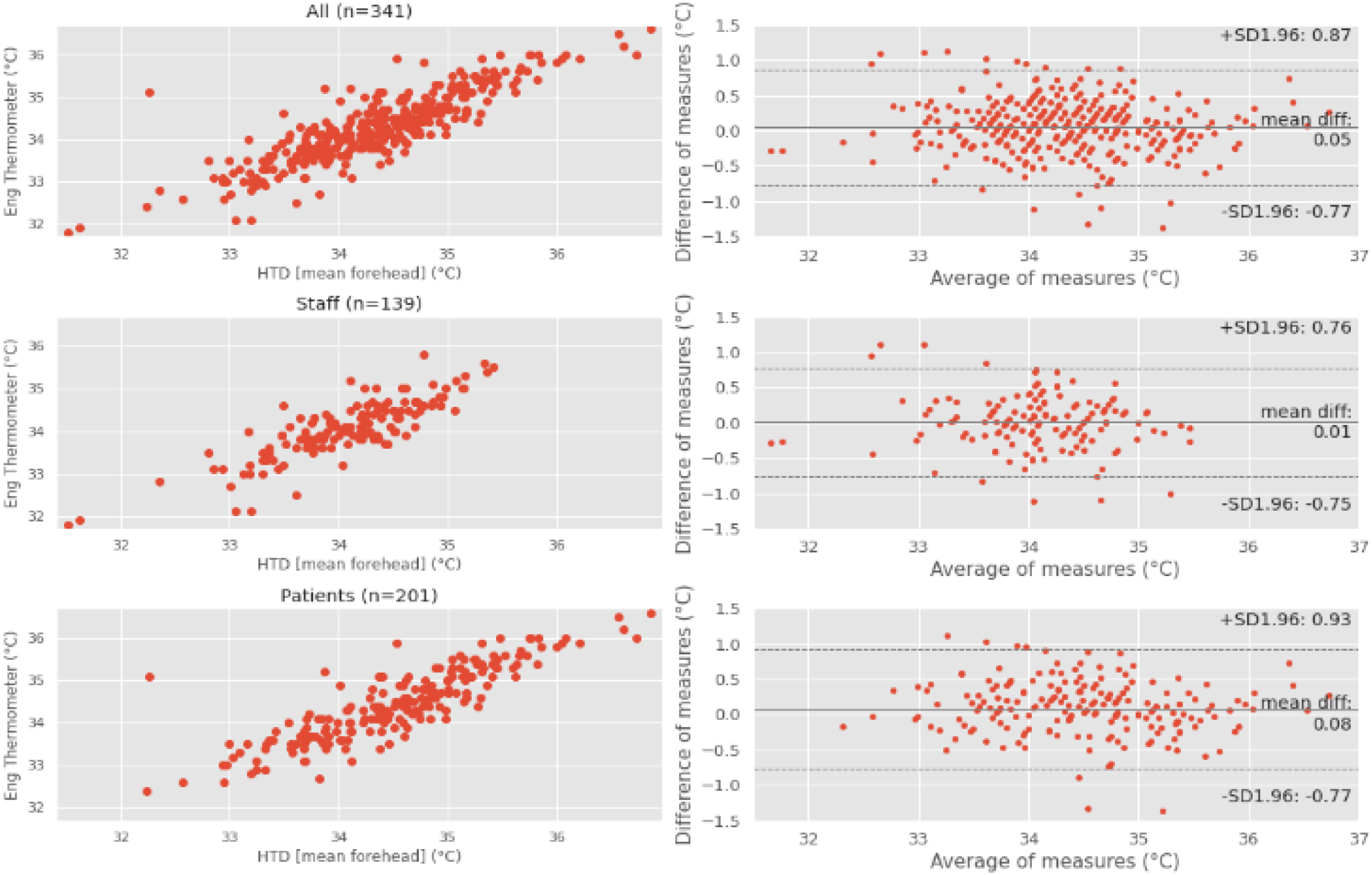
Comparison of Measurement of Mean Forehead Temperature using HTD camera and Engineering Thermometer

**Table 4:** Summary Statistics for Comparison of Measurement of Forehead Temperature by HTD Camera and Engineering Thermometer

| Set | Mean Error [$^{\circ}\text{C}$] | Standard Deviation [$^{\circ}\text{C}$] | RMS Error [$^{\circ}\text{C}$] | Correlation Coefficient |
| --- | --- | --- | --- | --- |
| All | 0.05 | 0.42 | 0.42 | 0.86 |
| Staff | 0.005 | 0.39 | 0.39 | 0.84 |
| Patients | 0.08 | 0.43 | 0.44 | 0.86 |

The mean error is negligible for the staff subset and small for the patients’ subset. The RMS error for the staff data set of 0.39°C is marginally bigger than the 0.27°C determined as a compound of errors presented in Table 3. However, this increase is not unexpected due to additional error in both measurements when measuring forehead temperatures compared to a static blackbody source. The correlation coefficients for all subsets are consistent and indicate a strong correlation between these measurements.

##### ii) Comparison HTD and COTS Forehead Thermometer

Figure 7 presents the comparison of the mean forehead skin temperature measurement using HTD camera and the COTS Forehead thermometer. Results for the three subsets are summarised in Table 5.

**Figure 7:**
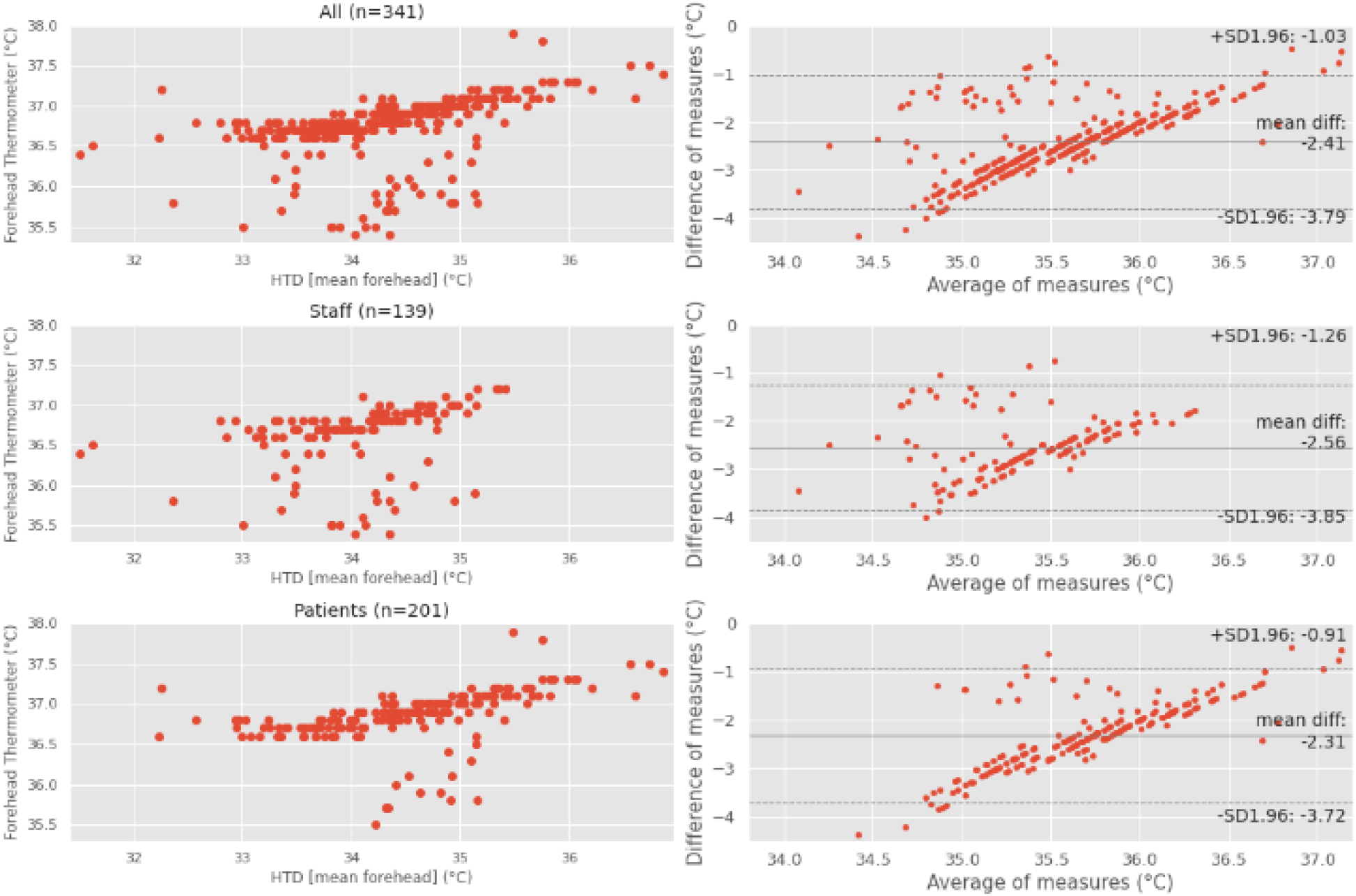
HTD Mean Forehead comparison to COTS Forehead thermometer

**Table 5:**
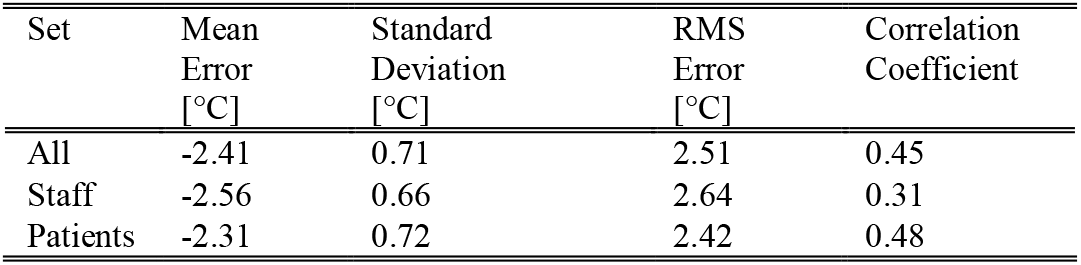
Summary Statistics for Comparison of Measurement of Forehead Temperature by HTD Camera and COTS Forehead Thermometer

| Set | Mean Error [°C] | Standard Deviation [°C] | RMS Error [°C] | Correlation Coefficient |
| --- | --- | --- | --- | --- |
| All | -2.41 | 0.71 | 2.51 | 0.45 |
| Staff | -2.56 | 0.66 | 2.64 | 0.31 |
| Patients | -2.31 | 0.72 | 2.42 | 0.48 |

The results show a weaker correlation than the Engineering thermometer. Similar to the results from measurement of the blackbody source presented in Section 1), the larger mean error is attributed to the core temperature estimation algorithm implemented by the COTS Forehead thermometer, which includes an offset from skin temperature to a predicted core temperature. The cause of the discrepancy between the correlation coefficients for the two data sets is uncertain.

### C. Relating HTD Measurements to Core Temperature as Measured by Clinical Reference Thermometers

Since the intended usage of the HTD camera is to estimate core temperature of the subject, a comparison of the temperatures measured to a core temperature measurement is required. Measurement of the tympanic temperature is the standard used in triage for measuring core temperature and identifying febrile patients in the Queen Elizabeth University Hospital. As such, measurements are compared to the tympanic temperature to imply any correlation with core temperature. It should be noted that the measurement of tympanic temperature to estimate core temperature is subject to inherent inaccuracy, and this must be considered when implying a correlation.

#### 1) Tympanic Temperatures by Demographic

Demographic distributions of measured tympanic temperatures for each demographic as collected in the trial are presented in Figure 1Figure 8. For the ethnic groups, White (*n*=358), Black/African/Caribbean/Black British (*n*=6), Asian/Asian British (*n*=20) and Other (*n*=8), the White demographic has the largest spread. The Black/African/Caribbean/Black British demographic has a lower median. However, due to limited sample sizes in groups other than White a reliable conclusion is not possible. Grouped by sex, the Female (*n*=225) sample has a higher median and smaller spread than the Male (*n*=167) sample. However the difference is minor and as such further analysis will be done on the sample as a whole. For age groups, ages 16-24 (*n*=50), 25-34 (*n*=117), 35-44 (*n*=62), 45-54 (*n*=62), 55-64 (*n*=36), 65-74 (*n*=33), 75-84 (*n*=26), and 85-94 (*n*=6) all indicate a similar median. Ages 35-44 and 65-74 have a large spread than the other groups. Age group 95+ had no samples.

**Figure 8:**
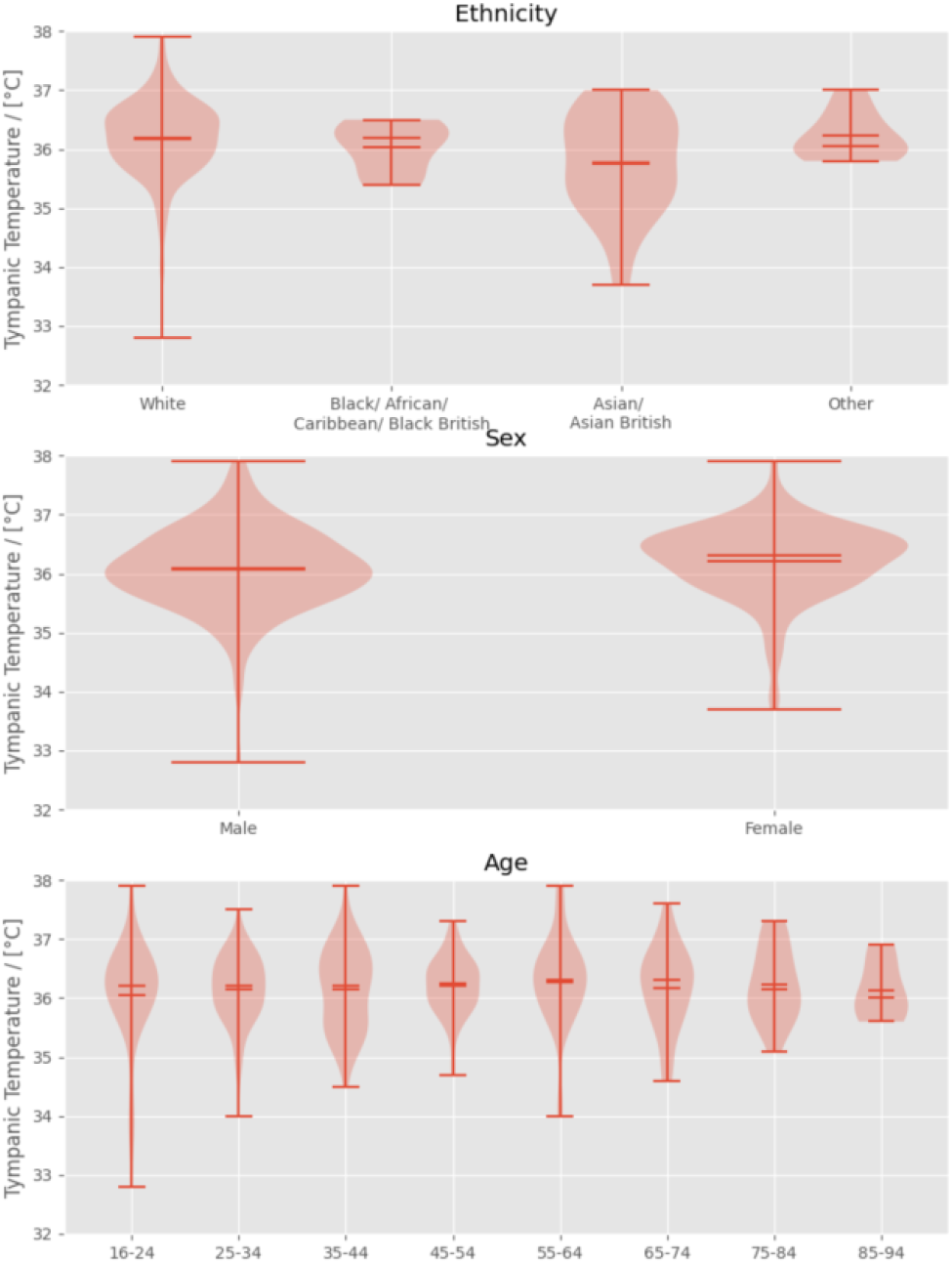
Tympanic temperatures by demographic

#### 2) Comparison of Measured Tympanic Temperature to Measured Forehead Skin Temperatures

Measured temperatures from the COTS Forehead and Engineering thermometer are compared to the tympanic thermometer and these results are presented in Figure 9. Comparisons of all skin temperature measurements and the tympanic temperature are summarised in Table 6.

**Figure 9:**
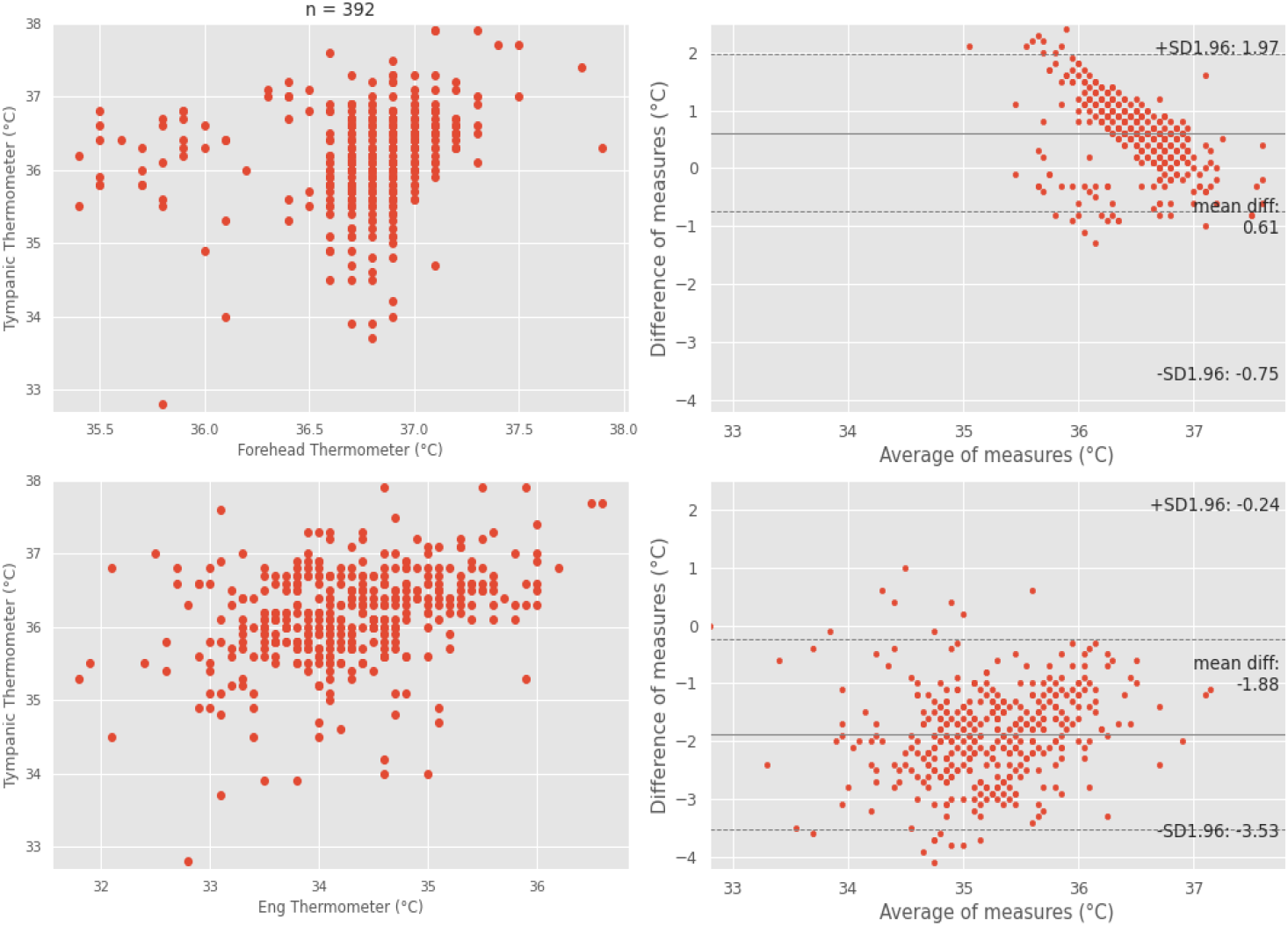
Comparison of COTS Forehead and Engineering Forehead Temperature Measurements to Tympanic Temperature Measurement

**Table 6:**
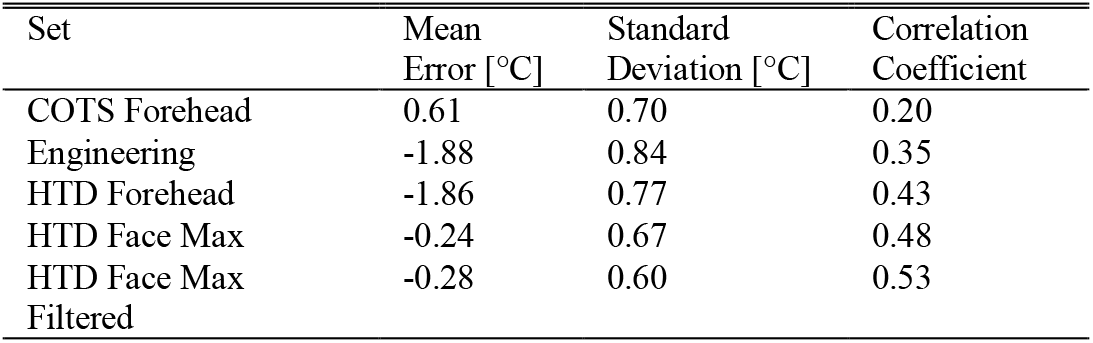
Summary Statistics for Comparison of Skin Temperature Measurements to Tympanic Temperature

| Set | Mean Error [°C] | Standard Deviation [°C] | Correlation Coefficient |
| --- | --- | --- | --- |
| COTS Forehead | 0.61 | 0.70 | 0.20 |
| Engineering | -1.88 | 0.84 | 0.35 |
| HTD Forehead | -1.86 | 0.77 | 0.43 |
| HTD Face Max | -0.24 | 0.67 | 0.48 |
| HTD Face Max Filtered | -0.28 | 0.60 | 0.53 |

In both instances, a weak correlation with tympanic temperature is observed, which implies a weak correlation to core temperature. This agrees with the literature which provides the consensus that skin temperature of the forehead is not a good indicator of core temperature [12].

Figure 10 presents equivalent results comparing the forehead measurement from the HTD system to that from the tympanic thermometer. The correlation is comparable if not stronger than that of the Engineering thermometer. The correlation is substantially stronger than the COTS Forehead thermometer. Assuming that tympanic temperature provides the strongest correlation to true core temperature, a core temperature estimation based on the forehead temperature measured by the HTD camera would yield a higher accuracy than the COTS Forehead thermometer used in this trial. A linear model used to predict core temperature based on HTD mean forehead temperature is shown in the bottom 2 plots of Figure 10, which when shown in a Bland-Altman analysis significantly reduces the observed bias between the two measurements, whilst also narrowing the 95% confidence interval.

**Figure 10:**
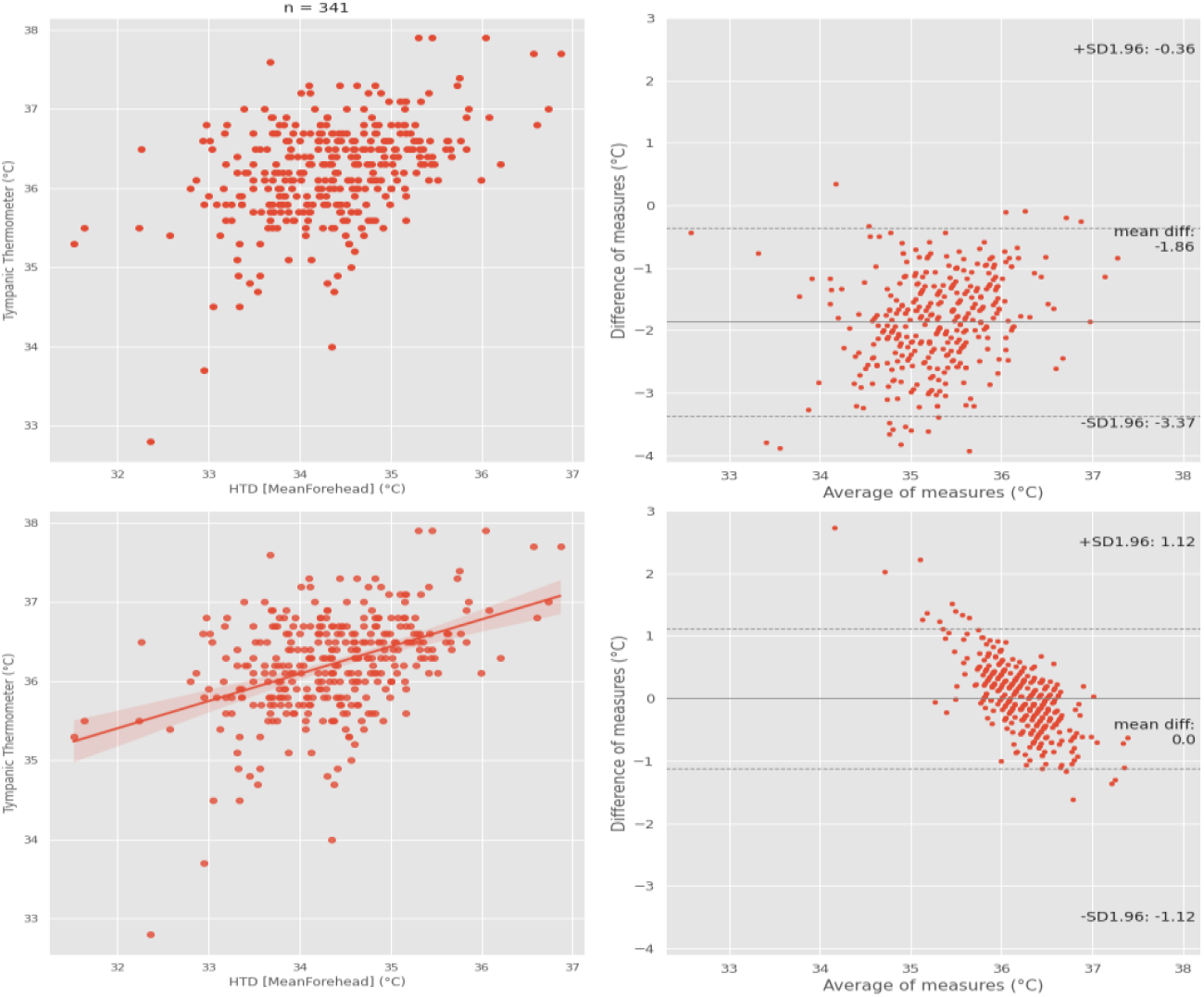
Comparison of HTD mean forehead to tympanic thermometer

#### 3) Comparison of Measured Tympanic Temperature to Measured Maximum Facial Temperature

More recent research focussing on both the maximum facial temperature and temperature of the inner canthus showed significantly stronger correlations between these temperatures and core temperature measured orally [13]. The maximum temperature from the face measured by the HTD system is compared to the tympanic thermometer. The results are shown in Figure 11.

**Figure 11:**
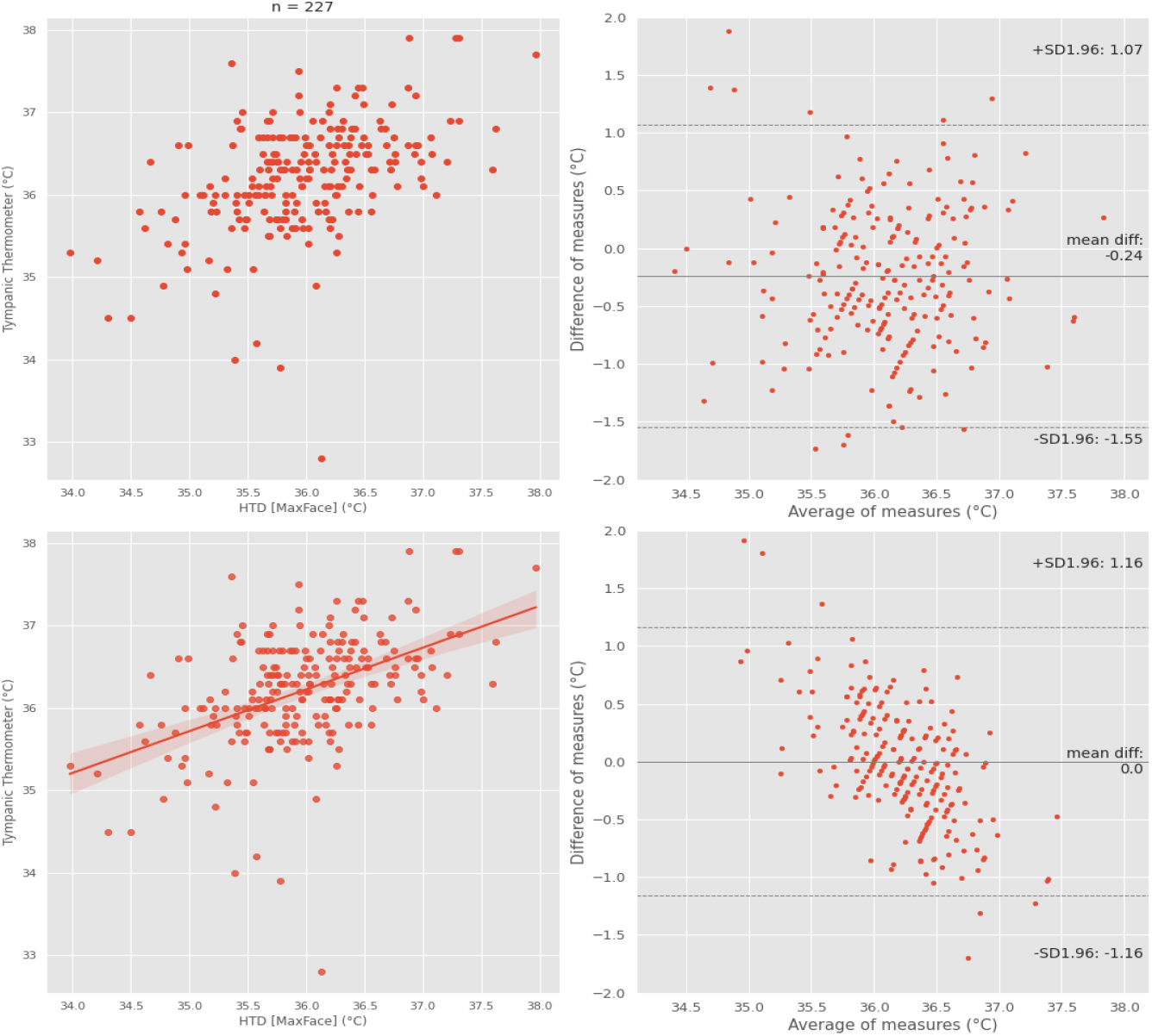
Comparison of HTD Max Face to Tympanic thermometer

The summary in Table 6 show a stronger correlation than that of all forehead temperature measurement. This is attributed to the maximum temperature of the face often reporting the temperature of the inner canthus. As such, using the same argument that tympanic temperature is best predictor of core temperature, it is implied that any core temperature estimation based on the maximum temperature from the face would produce a more accurate result than the methods based on skin temperature of the forehead.

Since the correlation is only moderate, a linear model is suggested for predicting core temperature from the maximum face temperature measured by HTD. The results of this model are shown in the Bland-Altman analysis in Figure 11.

The removal of four outliers further increases the correlation. Since these are outliers at the lower end of the distribution of tympanic temperatures, two arguments are presented for their exclusion: these points may have been incorrectly entered into the test application and the reliability of tympanic thermometer in predicting core temperature can vary from person to person, for example presence of ear wax in the ear canal.

Table 7 shows the results for error between tympanic temperature and HTD measurements with/without predictive model. The RMSE calculated for the linear model applied to the max face and compared to the tympanic temperature is equal to 0.50°C which is marginally larger than the 0.45°C error bound calculated as a compound of errors observed in Table 3. This suggests that the HTD camera can produce comparable results to the tympanic thermometer, while operating at range and without discomfort to the patient.

**Table 7:** Comparison between predictive models based on HTD Mean Forehead and Max Face against Tympanic Temperature

| Method | RMSE (Max Face, n = 223) | RMSE (Mean Forehead, n = 341) |
| --- | --- | --- |
| HTD (no change) | 0.71 | 2.01 |
| Rescaling based on tympanic distribution 1 | 0.67 | 0.67 |
| Linear model | 0.50 | 0.57 |

**Table 8:** Captured Data

| Field | Description |
| --- | --- |
| ID | Unique Identifier for Entry |
| Mean Selected Region | Mean of manually selected region |
| Max Selected Region | Max of manually selected region |
| Selection Region Bounding Box | Coordinates of manually selected region |
| Number of Faces Detected | Number of faces detected by Face Detection Algorithm |
| Mean Face Region | Mean of detected face region |
| Max Face Region | Max of detected face region |
| Face Region Bounding Box | Coordinates of detected face region |
| Face Max Point | Coordinate of maximum of detected face region |
| Engineering Thermometer Temperature | Temperature measured by the Engineering thermometer |
| COTS Forehead Thermometer Temperature | Temperature measured by COTS Forehead thermometer |
| Tympanic Temperature | Temperature measured by Tympanic thermometer |
| Age | Age in decile (16-24, 25-34, 35-44, 45-54, 55-64, 65-74, 75-84, 85+) |
| Sex | Sex |
| Ethnicity | Ethnicity (White, Mixed/multiple ethnic groups, Asian/Asian British, Black/Africa/Caribbean/Black British, Other) |

## V. Conclusions

When used in a clinical setting, the HTD camera achieved higher accuracy (*RMSEHTD*=0.15°*C*) compared to the Engineering IR thermometer (*RMSE*_*Engineering*_=0.22°*C*) when measuring a calibrated blackbody source set to typical skin temperature. This also demonstrates that the laboratory measured accuracy (*RMSE*_*HTD*_=0.1°*C*) is not significantly impacted when operated in a clinical setting. Measurement of the blackbody by the COTS Forehead thermometer shows an offset (*Mean Error*_*Forehead*_=2.06°*C*) from the blackbody set point, attributed to a core temperature estimation algorithm. The tympanic thermometer showed inferior accuracy (*RMSE*_*Tympanic*_=0.43°*C*) compared to the HTD camera, however direct comparison is difficult given difference in usage.

When measuring forehead skin temperature, comparing the HTD camera to the Engineering thermometer showed a strong correlation 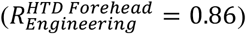 The mean error (*Mean Error*=0.05°*C*) and standard deviation (1*σ*=0.42°*C*) indicate that the HTD camera is capable of producing results commensurate to that of the Engineering thermometer, while being operated at distance.

Measurement and comparison of the HTD camera to the COTS Forehead thermometer show a weaker correlation 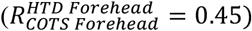. This weaker correlation is most likely attributable to the core temperature estimation algorithm included in the COTS Forehead thermometer and is observed in the mean error when comparing the HTD system and the COTS Forehead thermometer (*Mean Error*= −2.42°*C*).).

Comparison of the forehead skin temperature measurement by reference thermometers to the tympanic temperature measurement showed weak correlations 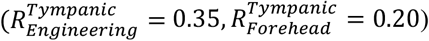 The HTD forehead measurement presented a marginally stronger correlation than both 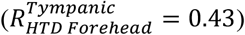 Although this correlation is only moderate, it is argued that measurement of the forehead by HTD is a better predictor of tympanic temperature and implies that it would be a more suitable predictor of core temperature than the COTS Forehead thermometer used in this trial. Furthermore, by comparison of the HTD maximum face temperature to the tympanic temperature, an improved correlation 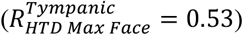 would indicate that any predictive model based on this HTD measurement could lead to a core temperature estimation with higher accuracy yield than all forehead skin temperature measurements while operated at a distance.

Applying a linear model to the maximum face temperature measured by HTD camera resulted in a predicted tympanic temperature 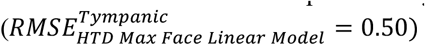 that is acceptable given the error inherent in the tympanic thermometer shown in measurement of the blackbody. Compared to the tympanic measurement, the HTD camera is operated at range, with zero discomfort for the patient and has the ability to be operated as passive screening in a clinical setting. The study demonstrates that the HTD could be applied in the clinical and non-clinical setting as a screening mechanism to detect citizens with raised temperature. This approach would enable high volume surveillance and identification of individuals that contribute to further spread of COVID-19. Deployment of the HTD system could be implemented as part of a screening tool to support measures to enhance public safety and confidence in areas of high throughput, such as airports, shopping centres or places of work.

## Data Availability

No external data was used in the preparation of the manuscript. 
All trials measurements and data is held by Thales in accordance with the requirements of the Trial and GDPR. All patient consent forms are held and archived by the Queen Elizabeth University Hospital in accordance with the requirements of the trial and GDPR. 

## Appendix A Data Filtering and Pre-processing

The data was divided into person and blackbody datasets. These were subsequently broken down into subsets containing valid data points for each measurement.

For the blackbody dataset, this included:

- The removal of 12 person entries that were incorrectly entered as blackbody measurements.
- The removal of 2 entries due to incorrect entry of both tympanic and forehead thermometer measurements.

For the blackbody HTD measurement subset this include:

- The removal of 11 entries due to incorrect physical setup of the system. These were captured on the first day of the trial and blackbody was positioned in a non-calibrated region of the image.
- The removal of 1 entry due to software error.
- The removal of 7 entries that are identified to be within the 15 minute settle period of the system.

For the person data set, this included:

- The removal of 1 entry due to incorrect entry of tympanic and forehead thermometer measurements.
- The removal of 2 entries due to software error.

The software error is attributed to a bug in the test application being used resulting in very large temperatures (in excess of 30000 degrees) and is not considered indicative of the accuracy of the system.

For the person HTD measurement subset, results from a further derived subset excluding 49 entries that were recorded on the initial day of the trial following prior to software changes in the test application are presented.

Additional metadata that was captured following the field trial includes the classification of the person as staff or patient, whether the person was wearing a mask and whether the person was wearing glasses. This data was appended through pre-processing the data.

## Appendix B Captured Data

Below is a complete list of metadata collected by the test application used in the trial.

## Acknowledgment

Thales would like to thank the team at Queen Elizabeth University Hospital (NHS Greater Glasgow and Clyde) for helping conduct this study under what are arguably some of the most challenging conditions seen in modern times.

Thales would also like to thank the West of Scotland Innovation Hub for development of the study and for project management support during the study.

This study was funded by Thales UK Ltd and Thales Digital Identity and Security Ltd, both subsidiaries of the Thales Group.

## Footnotes

1 It is expected as per Gauss-Markov theorem that the linear model is the best performing, the rescaling gives an example of intuitive model (the distribution of forehead temperature is transformed to match the one of tympanic temperature) and compares its performance to the best simple model

